# Effectiveness of booster BCG vaccination in preventing Covid-19 infection

**DOI:** 10.1101/2020.08.10.20172288

**Authors:** lradj Amirlak, Rifat Haddad, John Denis Hardy, Naief Suleiman Khaled, Michael H. Chung, Bardia Amirlak

## Abstract

**Introduction:** The evidence that BCG (bacille Calmette-Guerin) vaccine may increase the ability of the immune system to fight off pathogens other than tuberculosis has been studied in the past. This nonspecific immunity gained our interest, especially after initial reports of less cases in countries with universal BCG vaccination. In hopes of possible protective immunity, all staff of the Emirates International Hospital (United Arab Emirates) were offered a booster BCG vaccine in early March 2020. All the hospital staff were then tested for Covid-19 infection by the end of June 2020.

**Methodology:** We divided the subjects into two groups: booster vaccinated, versus unvaccinated. The rate of Covid-19 infection was compared between the groups. Criteria included all staff who were offered the vaccine.

**Results:** 71 subjects received the booster vaccination. This group had zero cases of positive COVID 19 infection. 209 subjects did not receive the vaccination, with 18 positive PCR confirmed COVID 19 cases The infection rate in the unvaccinated group was 8.6% versus zero in the booster vaccinated group. (Fisher’s exact test p-value=0.004).

**Conclusion:** Our findings demonstrated the potential effectiveness of the booster BCG vaccine, specifically the booster in preventing Covid-19 infections in an elevated-risk healthcare population.

## Introduction

BCG (bacille Calmette-Guerin), is a vaccine for M. tuberculosis infection (TB) both pulmonary and more so extra-pulmonary forms. It contains a live, attenuated form of Mycobacterium bovis strain and is only used in countries where the prevalence of TB is high. With initial reports of less cases and a lower death toll of Covid-19 in countries with universal childhood BCG vaccination, attention was turned to the non-specific effects (NSE) of this vaccine and how it may boost the immune response against Covid-19 ^1,2^. The Emirates International Hospital (EIH) functions as one of the centers for Covid-19 testing in the region as well as treating infected patients, putting the hospital staff at an elevated risk. In the first half of March 2020, at the upslope of the Covid-19 curve in the UAE, a decision was made by the EIH Covid-19 safety committee to offer abooster BCG (Serum Institute of India PVT.LTD. Hadaspar India) vaccination to the hospital staff. The prospect of increasing community infection, elevated risk to the hospital staff, and the available evidence about the BCG vaccine and potential non-specific effects against Covid-19 was taken into consideration. After thorough information about the vaccine and its prospects were provided to the hospital staff, voluntary vaccination was started. Out of 280 total staff, 71 received the vaccine in the span of two weeks. Compulsory government-mandated Covid-19 testing with PCR (CITOSWAB® nasopharyngeal swab) in the months of April, May, and June was done on all hospital staff. Testing was also was done in the setting of contact with positive patients and staff symptoms. The results of BCG immunity against Covid-19 infections was studied retrospectively.

## Methodology

After approval by the hospital review board and ethics committee, a retrospective cohort study of the entire hospital personnel was done and they were divided into two groups: those who received the BCG booster vaccine in March (group A), and those that did not (group B). Using a two-tailed Fisher’s exact test, the probability of testing positive in each group was compared.

## Results

Hospital staff were of Arabic, Indian, East Asian, African, and European origin. They were aged between 21 to 80 years and included clinical and non-clinical staff such as the receptionists, nurses, and physicians. By June 24th, 2020, 18 cases ofSARS-cov-2 infection were documented in the hospital staff (Figure 1). Group A included 71 individuals who received the BCG booster, and none tested positive for SARS-cov-2 infection. Group B, which included 209 non-booster vaccinated staff, had 18 positive PCR COVID-19 cases (Table 1), 13 of which were symptomatic and 5 had no symptoms. The infection rate was 18/209 or 8.6% in Group B, and zero in Group A. No reports of local or systemic complications was noted with the BCG booster vaccine group. The difference was statically significant, with a p-value of 0.004.

**Figure 1.**
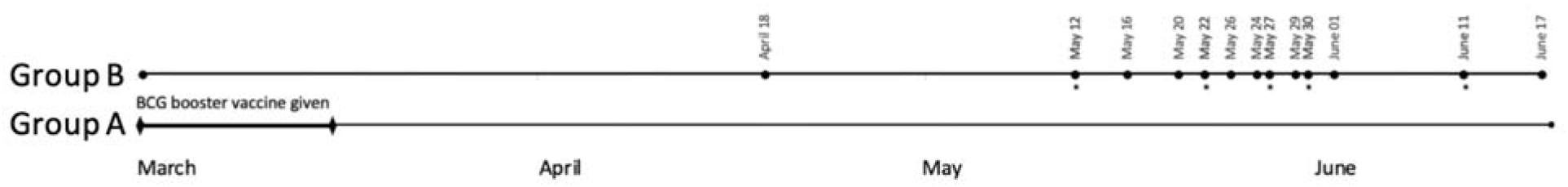
BCG booster vaccine was given to 71 subjects (Group A) in the first half of March 2020. 209 subjects opted out of the vaccine (Group B). Group A continued to have no positive cases through June 17, roughly 3 months after receiving the BCG booster. Group B’s first positive case was April 18. *Dates where two subjects tested positive included May 12, May 22, May 27, May 30, and June 11.

**Table 1.**
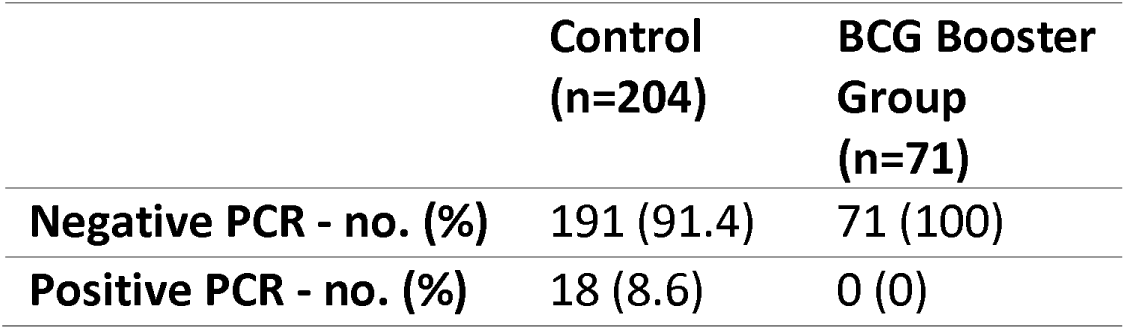
Outcomes in study cohort, P=0.005.

|  | Control<br>(n=204) | BCG Booster<br>Group<br>(n=71) |
| --- | --- | --- |
| Negative PCR - no. (%) | 191 (91.4) | 71 (100) |
| Positive PCR - no. (%) | 18 (8.6) | 0 (0) |

Both groups A and B had the BCG vaccine administered at birth, however no information was available about the exact brand of the BCG vaccine that were used. None of the Covid-19 positive staff had reported any high risk activity outside the hospital, infected household, or travel prior to their positive test. The positive Covid-19 subjects did not demonstrate any particular departmental and location distribution compared to the not infected. There was no collected data on hospital staff comorbidities and other medications used in either group.

## Discussion

Certain vaccines such as BCG, measles, and oral polio may have beneficial non-specific effects (NSE) beyond the pathogens they had been intended for ^3^. The epidemiological evidence that BCG may increase the ability of the immune system to fight off pathogens other than TB has been studied in the past, receiving mixed reviews initially and then a positive review by the World Health Organization ^4,5^. Such NSE’s are thought to be due to not only T cell–mediated adaptive immunity but also induction and enhancement of innate immunity^4,6^. It has been shown that the positive effect of the BCG vaccine given at birth or to children diminishes over time which likely indicates declining central memory immunity. It has been shown that a booster vaccine is needed to maintain the T cell response and antigen-specific central memory to battle TB infections years after the initial vaccine ^7,8^.

With lack of clinical data on the effectiveness of BCG vaccine in preventing Covid-19 infection, the decision to offer the vaccine to the hospital staff was based on available population based evidence, elevated risk of death and morbidity with the spread of Covid-19 amongst the staff, as well as proven safety of BCG vaccine over the years. This is a relatively small population sample who all work in the same environment with elevated levels of exposure to Covid-19 compared to the general population of the Al-Ain region of the UAE. None of the hospital staff who were booster BCG vaccinated tested positive with Covid-19 versus a 8.6% infection rate amongst those who were not recently vaccinated with a booster. There was similar universal personal protective equipment policy throughout the hospital with no specific pattern of infection correlating to the location of work in the hospital. Thus we assumed that any discrepancy of exposure level is minimized between the two groups.

Despite population-based findings on the protective role of BCG vaccination on Covid-19 infection ^1,2,9^ a recent study by Hamiel et al in young adults found that childhood vaccination may not have a significant protective effect in younger people ^10^. All our 280 hospital staff (Group A and B) were vaccinated with BCG at birth. The lack of Covid-19 infection in our recently vaccinated versus the high rate of infection in the non-recently vaccinated group, suggest stronger protective effect of booster BCG vaccination compared to birth vaccination. The effect of such NSE immunity are poorly understood and we do not know how long the presumed immunity may last.

The limitations of our study are several, such as lack of clear understanding and documentation of any confounding factors between the two groups that could have influenced the transmission and infection rate, sample size, and discrepancy between the number of subjects in the two group. Staff comorbidity as well as any over the counter supplements that the staff could have taken in attempts to prevent Covid-19 infection was not documented. Despite the limitations of this study, we feel that our findings of 8.6% versus zero percent infection rate is significant enough to suggest the promising effectiveness of an up-to-date BCG booster vaccine in prevention of Covid-19 infection and therefore warrants reporting.

It is important to note that despite the statistically significant clinical evidence presented here, disregarding the study limitations and recommending mass BCG vaccination before prospective trials is an overreach and could create a false and dangerous sense of confidence in vaccine recipients. The health authority in many countries have recommended a booster BCG vaccine to prevent TB infection^11^. Some physicians may consider providing voluntary booster BCG vaccination, especially in areas of substantial Covid-19 outbreak in which tuberculosis is already prevalent. If such decision is made, it should be in accordance to the local ethical medical practice principles, with full disclosure of treatment and preventative options, short term and long term side effects and without misleading or deceptive promises made to patients. Adverse effects of BCG vaccine are rare local reactions that are temporary, and extremely rare serious complication of disseminated BCG infection. BCG vaccination should not be given during pregnancy, to persons who are immunosuppressed, or who are likely to become immunocompromised ^12^.

## Conclusion

Our findings are the first and only one to date to demonstrate possible NSE immunity to SARS-cov-2 infection in a clinical setting. Prospective studies with vaccination of high risk health care workers can further clarify the relationship of booster BCG vaccine and the level of immunity it provides.

## Data Availability

Data available if requested.

## Notes

### Competing Interest Statement

The authors have declared no competing interest.

### Funding Statement

No funding to be reported.

### Author Declarations

Emirates International Hospital review board and ethics committee

## References

1. Berg MK, Yu Q, Salvador CE, Melani I, Kitayama S. Mandated Bacillus Calmette-Guérin (BCG) vaccination predicts flattened curves for the spread of COVID-19. *medRxiv*. 2020:2020.2004.2005.20054163.

2. Miller A, Reandelar MJ, Fasciglione K, Roumenova V, Li Y, Otazu GH. Correlation between universal BCG vaccination policy and reduced morbidity and mortality for COVID-19: an epidemiological study. *medRxiv*. 2020:2020.2003.2024.20042937.

3. Higgins J, Soares-Weiser K, Reingold A. Systematic review of the non-specific effects of BCG, DTP and measles containing vaccines. Report to WHO 2014.

4. Leentjens J, Kox M, Stokman R, et al. BCG Vaccination Enhances the Immunogenicity of Subsequent Influenza Vaccination in Healthy Volunteers: A Randomized, Placebo-Controlled Pilot Study. J Infect Dis. 2015;212(12):1930–1938.

5. Higgins JP, Soares-Weiser K, Lopez-Lopez JA, et al. Association of BCG, DTP, and measles containing vaccines with childhood mortality: systematic review. BMJ. 2016;355:i5170.

6. Netea MG, van Crevel R. BCG-induced protection: effects on innate immune memory. Semin Immunol. 2014;26(6):512–517.

7. Gupta N, Garg S, Vedi S, Kunimoto DY, Kumar R, Agrawal B. Future Path Toward TB Vaccine Development: Boosting BCG or Reeducating by a New Subunit Vaccine. Front Immunol. 2018;9:2371.

8. Whittaker E, Nicol MP, Zar HJ, Tena-Coki NG, Kampmann B. Age-related waning of immune responses to BCG in healthy children supports the need for a booster dose of BCG in TB endemic countries. Sci Rep. 2018;8(1):15309.

9. Escobar LE, Molina-Cruz A, Barillas-Mury C. BCG vaccine protection from severe coronavirus disease 2019 (COVID-19). Proceedings of the National Academy of Sciences. 2020:202008410.

10. Hamiel U, Kozer E, Youngster I. SARS-CoV-2 Rates in BCG-Vaccinated and Unvaccinated Young Adults. JAMA. 2020.

11. Zwerling A, Behr MA, Verma A, Brewer TF, Menzies D, Pai M. The BCG World Atlas: a database of global BCG vaccination policies and practices. PLoS Med. 2011;8(3):e1001012.

12. BCG vaccines: WHO position paper-February 2018. Geneva: World Health organization; 2018.

